# Performance evaluation of the BD SARS-CoV-2 reagents for the BD MAX™ system

**DOI:** 10.1101/2021.05.12.21257120

**Authors:** Karen Yanson, William Laviers, Lori Neely, Elizabeth Lockamy, Luis Carlos Castillo-Hernandez, Christopher Oldfied, Ronald Ackerman, Jamie Ackerman, Daniel A Ortiz, Sixto Pacheco, Patricia J Simner, Stephen Young, Erin McElvania, Charles K Cooper

**Affiliations:** Becton, Dickinson and Company, BD Life Sciences – Integrated Diagnostic Solutions, 7 Loveton Circle, Sparks, MD, USA; CTMD Research, 2328 S. Congress Ave., STE 1-E, Palm Springs, FL, USA; Fellows Research Alliance, Inc., 1 Oglethorpe Professional Blvd, Suite 204, Savannah, GA, USA; Comprehensive Clinical Research, LLC, 603 Village Blvd, Suite 301, West Palm, FL, USA; William Beaumont Hospital, 3601 W 13 Mile Rd, Royal Oak, MI, USA; BioCollections Worldwide, Inc., 5735 NE 2nd Avenue, Miami, FL, USA; Johns Hopkins University School of Medicine, Department of Pathology, Division of Medical Microbiology, Meyer B1-125D, 600 N. Wolfe Street, Baltimore, MD, USA; Tricore Reference Laboratory, 1001 Woodward Place, N.E., Albuquerque, NM, USA; NorthShore University Health System, Department of Pathology and Laboratory Medicine, 2650 Ridge Avenue, Evanston, IL, USA

## Abstract

**Background:** The RT-qPCR assay for detecting SARS-CoV-2 virus is the favorable approach to test suspected COVID-19 cases. However, discordant results can occur when two or more assays are compared. Variability in analytical sensitivities between assays, among other factors, may account for these differences in reporting.

**Methods:** The limits of detection (LOD) for the BD SARS-CoV-2 Reagents for BD MAX™ System (“MAX SARS-CoV-2 assay”), the Biomerieux BioFire^®^ Respiratory Panel 2.1 (“BioFire SARS-CoV-2 assay”), the Roche cobas SARS-CoV-2 assay (“cobas SARS-CoV-2 assay”), and the Hologic Aptima® SARS-CoV-2 assay Panther® (“Aptima SARS-CoV-2 assay”) RT-qPCR systems were determined using a total of 84 contrived nasopharyngeal specimens with seven target levels for each comparator. The positive and negative percent agreement (PPA and NPA, respectively) for the MAX SARS-CoV-2 assay were compared to the Aptima SARS-CoV-2 assay in a post-market clinical study utilizing 708 paired nasopharyngeal specimens collected from suspected COVID-19 cases. Discordant results were further tested by the cobas and BioFire SARS-CoV-2 assays.

**Results:** The measured LOD for the MAX SARS-CoV-2 assay (251 copies/mL) was comparable to the cobas SARS-CoV-2 assay (298 copies/mL) and the BioFire SARS-CoV-2 assay (302 copies/mL); the Aptima SARS-CoV-2 assay had a LOD of 612 copies/mL. The MAX SARS-CoV-2 assay had a PPA of 100% (95%CI: [97.3%-100.0%]) and a NPA of 96.7% (95%CI: [94.9%-97.9%]) when compared to the Aptima SARS-CoV-2 assay.

**Conclusions:** The MAX SARS-CoV-2 assay exhibited a high analytical sensitivity and specificity for SARS-CoV-2 detection. The clinical performance of the MAX SARS-CoV-2 assay agreed with another sensitive EUA cleared assay.

## INTRODUCTION

Since December 2019, when a cluster of cases was first reported in Wuhan, China, the COVID-19 pandemic, caused by the SARS-CoV-2 virus, has been a major public health crisis, globally.(1) As of mid-May 2021, more than 160 million cases and 3.3 million deaths have been identified, worldwide, with more than 32.8 million cases and 582 thousand deaths in the U.S. alone.(2) Rapid transmission and lack of treatment make it difficult to mitigate the pandemic.(3) Isolating suspected patients and executing effective contact tracing is critical for managing the spread of the disease.(4) Diagnosis of COVID-19, through accurate detection of SARS-CoV-2 is the first step in guiding healthcare providers to triage patients, determine the treatment plan, and quarantine suspected contacts.

Diagnostic testing methodology for SARS-CoV-2 detection has been rapidly implemented in response to the pandemic.(5) The molecular or nucleic acid testing using the real-time, reverse transcriptase-quantitative polymerase chain reaction (RT-qPCR) assay has been a standard way to detect SARS-CoV-2 and diagnose COVID-19.(6) RT-qPCR-based assays are effective for SARS-CoV-2 nucleic acid detection in upper respiratory specimens collected through swab sampling.(7, 8) This approach generally exhibits advantages in sensitivity and specificity in specimens collected from nasopharyngeal, oropharyngeal, mid-turbinate nasal, or anterior nasal swabs. Although the overall turnaround time has been lengthy traditionally, the implementation of molecular testing on automated platforms has helped to ensure better turn-around-times for RT-qPCR testing while maintaining a high level of performance for detection of SARS-CoV-2.

In response to the COVID-19 pandemic, World Health Organization (WHO) and the U.S. Food and Drug Administration (FDA) issued emergency use authorization (EUA) for the development of *in vitro* diagnostic assays.(5, 9) Several manufactures have developed RT-qPCR platforms for SARS-CoV-2 testing. Most of them are intended for testing nasal, nasopharyngeal, and oropharyngeal swab samples collected from individuals suspected of having COVID-19. The BD SARS-CoV-2 Reagents for BD MAX™ System (“MAX SARS-CoV-2 assay;” Becton, Dickinson and Company; BD Life Sciences – Integrated Diagnostics Solutions, Sparks, MD, USA) utilizes multiplexed primers and probes that are designed to amplify two unique regions of the SARS-CoV-2 nucleocapsid (N) gene, N1 and N2, and the human ribonucleases P (RNase P) gene and received FDA EUA on April 8, 2020.(10) Following the initial EUA, two modifications were authorized (on March 10, 2021) by the FDA for the assay: (a) an increase to the cutoff for the N2 channel, (b) an improvement to the probe chemistry to reduce the background fluorescence.(11)

The objective here was to assess the performance of the modified MAX SARS-CoV-2 assay for detection of SARS-CoV-2 in nasopharyngeal specimens collected consecutively from individuals suspected of COVID-19. The analytical sensitivity was first determined for the MAX SARS-CoV-2 assay and three other commercial SARS-CoV-2 RT-qPCR assays: the Biomerieux BioFire^®^ Respiratory Panel 2.1 (“BioFire SARS-CoV-2 assay;” Biomerieux, BioFire Diagnostics, Salt Lake City, UT, USA), the Roche cobas SARS-CoV-2 assay (“cobas SARS-CoV-2 assay;” Roche Diagnostics, Indianapolis, IN, USA), and the Hologic Aptima® SARS-CoV-2 assay Panther® System (“Aptima SARS-CoV-2 assay;” Hologic, Marlborough, MA, USA). The clinical performance of the MAX SARS-CoV-2 assay was further examined by determining the positive percent and negative percent agreements (PPA and NPA, respectively) with the Aptima SARS-CoV-2 assay. The utilization of multiple assays here facilitated comprehensive discordant testing in the absence of an established clinical reference standard SARS-CoV-2 PCR-based assay.

## METHODS AND MATERIALS

### Specimens and assays

The first study compared the analytical sensitivity of the MAX SARS-CoV-2 assay, the BioFire SARS-CoV-2 assay, the cobas SARS-CoV-2 assay, and the Aptima SARS-CoV-2 assay. A total of 84 contrived nasopharyngeal specimens were prepared for each commercial assay. The specimens were diluted in universal viral transport media to generate a panel consisting replicates of six concentrations (22, 67, 200, 600, 1800, and 5400 copies/mL) for each assay. An additional negative control level was also prepared for each panel.

The second study involved post-market clinical testing and involved 1,376 specimens from four collection sites in the U.S. (Table S1). The specimens included prospective as well as consecutively collected remnant nasopharyngeal swabs from symptomatic patients suspected of COVID-19 by their healthcare providers. There were 64 specimens that had an Aptima SARS-CoV-2 result but were not tested on MAX SARS-CoV-2. There were also 288 specimens that were enrolled but were not tested on either Aptima SARS-CoV-2 or MAX SARS-CoV-2 since the positive target goal was attained. Overall, 708 paired specimens were utilized for testing and analysis. Demographic information for compliant specimens with reportable results is shown in Table S2. The study protocol was approved by the Advarra Institutional Review Board and de-identified specimens from collection sites were used for testing. Written, informed consent was obtained prior to any trial-related procedures. This study was conducted according to the principles set forth by the Declaration of Helsinki and Good Clinical Practice.

### Data analysis

The analytical sensitivity values for the four assays were determined by calculating the limit of detection (LOD) using probit regression analysis. The point estimate for LOD is the lowest detectable concentration of SARS-CoV-2 at which approximately 95% of all (true positive) replicates test positive. Goodness-of-fit test was performed using Pearson and deviance correlation methods. Only data following normality or having at least two functional data points from a comparator yielded an appropriate statistical fit.

For the post-market clinical study, the primary outcome measures were PPA and NPA point estimates (with calculated 95% confidence intervals [95% CI] using the Wilson score method) for the MAX SARS-CoV-2 assay, compared to the reference assay, Aptima SARS-CoV-2. Cohen’s kappa coefficient was utilized to gauge the agreement between two raters (reference versus index test) to classify results into mutually exclusive categories. Κ=(P_o_^-P^e)/1-P_e_ (<0, 0, and >0 indicate agreements worse than, no better or worse than, and better than that expected by chance). Acceptance criteria for the MAX SARS-CoV-2 assay for FDA-EUA clearance for SARS-CoV-2 were ≥95% for both PPA and NPA.(9) Only compliant and reportable results for both MAX SARS-CoV-2 and comparator assays were included. This article was prepared according to STARD guidelines for diagnostic accuracy studies reporting.(12) Data will be made publicly available upon publication and upon request for peer review.

## RESULTS

The MAX SARS-CoV-2 assay was subjected to a series of validations to determine the impact (if any) on analytical sensitivity and specificity resulting from the cutoff change on the N2 channel and the modification to the probe chemistry.(10) As shown in Table 1, the LOD of the MAX SARS-CoV-2 assay was obtained and compared to three other commercially available SARS-CoV-2 RT-qPCR assays, specifically, the BioFire SARS-CoV-2, the cobas SARS-CoV-2, and the Aptima SARS-CoV-2 assays. From a total of 84 contrived nasopharyngeal specimens with seven target levels, the MAX SARS-CoV-2 assay had the lowest LOD (251 copies/mL), but was comparable to cobas SARS-CoV-2 (298 copies/mL for Target 2) and BioFire SARS-CoV-2 (302 copies/mL) assays. The Aptima SARS-CoV-2 assay showed the highest LOD (612 copies/mL), but was within a 2-fold concentration range of the other assays.

**Table 1.**
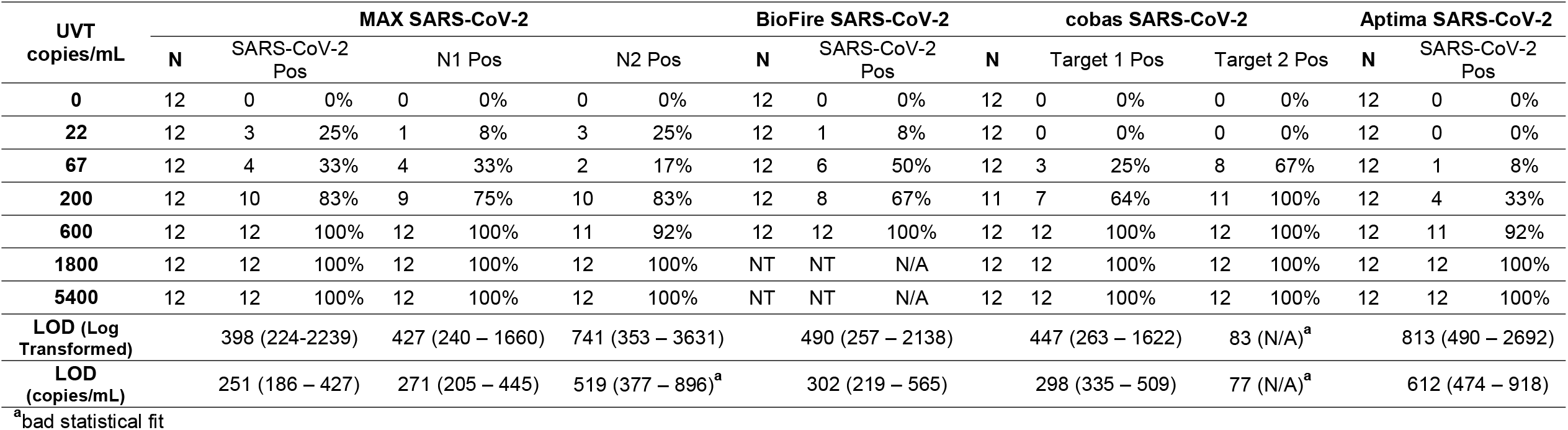
Limit of detection (LOD) of MAX SARS-CoV-2, BioFire SARS-CoV-2, cobas SARS-CoV-2 and Aptima SARS-CoV-2.

In the clinical evaluation study, a total of 708 specimens tested were included for paired analysis. Among all analyzed samples, 138 were positive by both MAX SARS-CoV-2 and Aptima SARS-CoV-2 assays, while 551 tested negative by both assays. Therefore, MAX SARS-CoV-2 testing resulted in a PPA of 100% (95%CI: [97.3%-100.0%]) and a NPA of 96.7% (95%CI: [94.9%-97.9%]), when compared to the Aptima SARS-CoV-2 assay (Table 2). Discordant results were observed from 19 specimens that were positive with the MAX SARS-CoV-2 assay but negative by the Aptima SARS-CoV-2 assay (Table 3). Among these, 5 specimens were N1 positive/N2 positive, 11 specimens were N1 positive/N2 negative, and 3 specimens were N1 negative/N2 positive. The BioFire SARS-CoV-2 and the cobas SARS-CoV-2 assays were utilized for discrepancy testing. Of the 19 discordant specimens, 4 tested positive by the cobas SARS-CoV-2 assay and 5 were positive by the BioFire SARS-CoV-2 assay. One of the specimens did not generate a reportable result from either the cobas SARS-CoV-2 assay or the BioFire SARS-CoV-2 assay. Five (5) of the 19 discordant specimens did not have sufficient volume for BioFire SARS-CoV-2 testing and 5 did not yield valid results from the cobas SARS-CoV-2 assay due to a low volume error. Overall, 7 of 19 MAX SARS-CoV-2-positive specimens were also positive in discordant testing. Further analysis revealed that only one of the specimen results corresponded to the MAX SARS-CoV-2 Ct values less than 30 (specimen ID #9 in Table 3); the other results were either at, or close to, the LOD for the MAX SARS-CoV-2 assay.

**Table 2.**
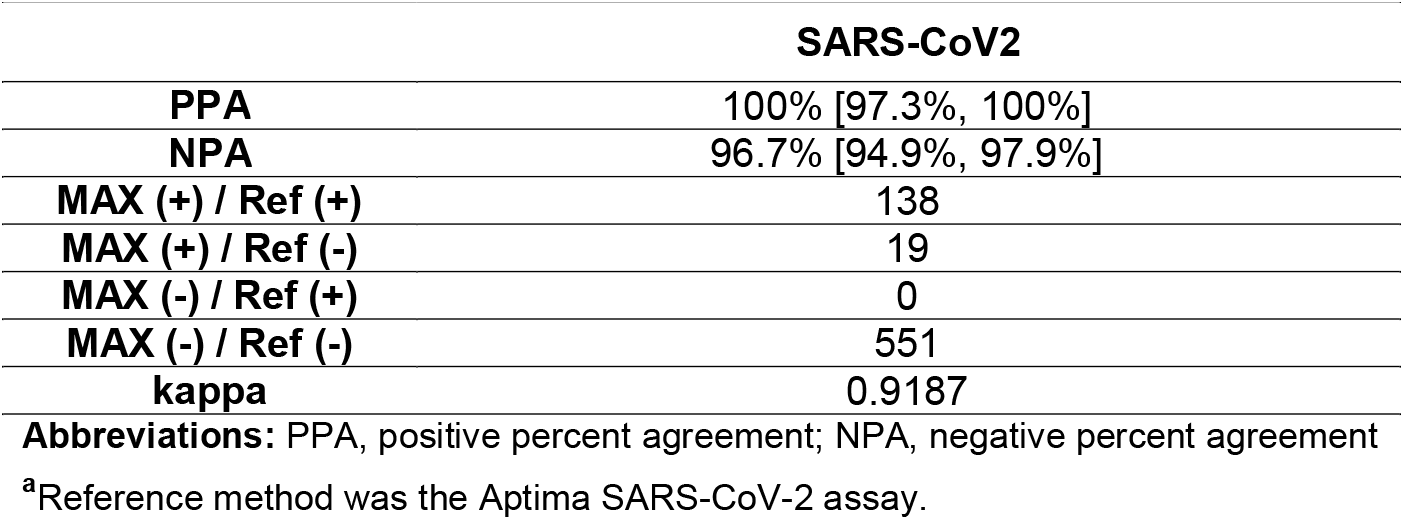
Performance of the MAX SARS-CoV-2 assay for detection of SARS-CoV-2 compared to reference.^**a**^

**Table 3.** List of MAX SARS-CoV-2 (+) / Aptima SARS-CoV-2 (-) specimens.

| Specimen ID | MAX N1 Ct1 | MAX N2 Ct | MAX RNaseP Ct | cobas Target 1 | cobas Target 2 | BioFire |
| --- | --- | --- | --- | --- | --- | --- |
| 1 | 37.1 | -1 | 24.7 | + (33.86) | + (35.25) | + |
| 2 | 34.9 | 33.2 | 22.5 | - | + (36.99) | + |
| 3 | 34.9 | -1 | 21 | - | + (35.27) | QNS |
| 4 | 37.9 | -1 | 23.4 | - | + (37.02) | QNS |
| 5 | -1 | 35 | 22.3 | INV | INV | + |
| 6 | 35.3 | 34.4 | 26.4 | INV | INV | + |
| 7 | 37.5 | -1 | 24 | - | - | + |
| 8 | 40.2 | -1 | 22.8 | - | - | QNS |
| 9<sup>a</sup> | 22.9 | 24.5 | 19.7 | - | - | QNS |
| 10 | 38.8 | -1 | 22 | INV | INV | - |
| 11 | 37.4 | -1 | 26.2 | INV | INV | - |
| 12 | 33.3 | 33.2 | 28.1 | - | - | - |
| 13 | 39.4 | -1 | 25.1 | - | - | - |
| 14 | 35 | -1 | 25 | - | - | - |
| 15 | 33.8 | -1 | 26.8 | - | - | - |
| 16 | 31.2 | -1 | 29.1 | - | - | - |
| 17 | -1 | 35.9 | 26.8 | - | - | - |
| 18 | -1 | 35.5 | 19.4 | - | - | - |
| 19 | 32.9 | 33.6 | 22.5 | INV | INV | QNS |

**Abbreviations:** INV, invalid value; QNS, quantity not sufficient for testing
<sup>a</sup>With the exception of sample 9, all discordant samples exhibit Ct values at or near the limit of detection on the MAX SARS-CoV-2 system.

## DISCUSSION

On March 10, 2021, the FDA provided EUA for the modified MAX SARS-CoV-2 RT-qPCR assay, which eliminated the condition of follow-up testing for a MAX SARS-CoV-2 assay presumptive positive.(11) Here, the MAX SARS-CoV-2 assay showed a relatively low LOD (251 copies/mL) when compared with three other RT-qPCR EUA assays. The clinical study, incorporating 708 real-world specimens, resulted in 100% PPA and 96.7% NPA performance values for MAX SARS-CoV-2 when compared to the reference assay, and met FDA EUA acceptance criteria of greater than 95% for PPA and NPA. This work demonstrates that the MAX SARS-CoV-2 assay has excellent analytical sensitivity and clinical agreement for detection of SARS-CoV-2.

In this study, the MAX SARS-CoV-2 assay was first compared to the Aptima SARS-CoV-2 assay. Nineteen (19) discordant results occurred from 708 paired specimens. Upon discordant method testing, five specimens were positive by at least one additional RT-qPCR assay and two additional specimens were positive for cobas SARS-CoV-2 Target 2 (sarbecovirus). Several factors, including primer design, type of polymerase employed, reaction conditions, and template purity could all impact the analytical sensitivity of PCR.(13) In the absence of a consensus clinical reference standard for detection of SARS-CoV-2 via nucleic acid amplification, it is difficult to adjudicate, with certainty which of the remaining 14 positive specimens by the MAX SARS-CoV-2 assay were truly positive. The benchmarking data here demonstrated that the MAX SARS-CoV-2 assay had the lowest LOD of the four assays tested. Interestingly, the genomic organization of SARS-CoV-2 viral RNA includes the open reading frame 1 (ORF1) and several subgenomic regions encoding the structural proteins, such as spike protein (S), envelope (E), membrane protein (M), and nucleocapsid (N) (Figure S1A). Among these subgenomic regions, the N gene expresses the most abundant transcript, and could provide a higher starting amount of template, giving the MAX SARS-CoV-2 assay a lower apparent LOD.(14) In addition, the larger input volume associated with the MAX SARS-CoV-2 assay could facilitate a lower LOD by further providing more template from which to amplify (Figure S1B).(10)

As the genetic mutations of SARS-CoV-2 virus quickly evolve to account for several novel viral variants that exhibit higher transmission and mortality rate, whether the current RT-qPCR assays can accurately detect the virus becomes the topic of interest.(15, 16) If genetic mutations occur within the target regions of the SARS-CoV-2 primer, proper primer and probe binding may be affected and fail to detect the presence of the virus, correctly.(17) To date, multiple mutations have been mapped out and most of them are identified at the ORF1 region, followed by S, N, M, and E gene (Figure S1A).(18) It is important to note that the mutations occur on S gene often result in changes of viral transmission. The specificity of the N1 and N2 primer sets used by MAX SARS-CoV-2 assay was determined using an *in silico* approach to compare with all 329,434 available SARS-CoV-2 sequences in the GISAID database as of January 21, 2021. Alignments against the N gene showed that both N1 and N2 primer/probe sets are a perfect match to 93.8% of sequences in the database, 96.8% of the sequences are a perfect match to the N1 primer set region, and 97.0% are a perfect match to the N2 primer set region. In total, 99.9% are a perfect match to either the N1 or the N2 region primer set. Additionally, N1 and N2 primers showed no significant combined homologies with human genome regions, other coronaviruses, or human microflora that would predict potential false positive RT-qPCR results. On the other hand, with Aptima and cobas SARS-CoV-2 assays recognizing ORF1 gene and BioFire SARS-CoV-2 assay targeting the S and M genes, the detection of mutated SARS-CoV-2 may be missed (Figure S1A and B).(10, 19-21)

Although the RT-qPCR testing method is a highly sensitive and favorable approach to detect SARS-CoV-2 virus, the positive results do not rule out bacterial infection or co-infection with other viruses. On the other hand, negative results do not preclude SARS-CoV-2 infection, and should be considered in conjunction with clinical observations, medical history, and epidemiological information. Furthermore, all factors mentioned in the previous paragraphs could potentially affect the detection ability of the system and one hundred percent agreement between assays should not be expected. While the false negative result creates a major public health concern for mitigating the pandemic, the false positive may produce unnecessary stress on workforce management and treatment planning for other diseases.(22-24) Without the gold standard test available to which the results of the RT-qPCR can be compared, combining the clinical presentations with the test results may better diagnose the infectious status of COVID-19.(23, 25, 26)

## Limitations

Not all discordant specimens were tested by two other assays, cobas SARS-CoV-2 and BioFire SARS-CoV-2, due to the specimen volume available.

## Conclusions

MAX SARS-CoV-2 assay has two targets specific for the N gene of the SARS-CoV-2 virus that contributes to the high analytical sensitivity and specificity in detecting the virus. MAX SARS-CoV-2 assay exhibited strong clinical agreement to another EUA assay with more positives detected as confirmed by the discordant methodology.

## Data Availability

Data will be available upon request.

## ACKNOWLEDGEMENTS

We thank Yu-Chih Lin, Ph.D. (Becton, Dickinson and Company, BD Life Sciences – Integrated Diagnostic Solutions) for the input on the content of this manuscript and editorial assistance. We thank Stanley Chao and Yongqiang Zhang (Becton, Dickinson and Company, BD Life Sciences – Integrated Diagnostic Solutions) for statistical support. The individuals acknowledged here have no additional funding or additional compensation to disclose. We are grateful to the study participants who allowed this work to be performed.

## AUTHOR CONTRIBUTIONS

All authors contributed to the interpretation of the data, critically revised the manuscript for important intellectual content, approved the final version to be published, and agree to be accountable for all aspects of the work.

---

**Supplemental Table 1.** Enrollment and compliance summary.

| **Collection Site** | **Total #**<br>**Enrolled** | **# of Compliant**<br>**Specimens** | **Reportable Aptima and**<br>**MAX Result<sup>a</sup>** |
| --- | --- | --- | --- |
| A | 248 | 244 | 110 |
| B | 450 | 445 | 237 |
| C | 150 | 141 | 130 |
| D | 528 | 231 | 231 |
| Overall | 1376 | 1061 | 708 |

<sup>a</sup>There were 64 specimens that had an Aptima SARS-CoV-2 result but were not tested on MAX SARS-CoV-2. There were also 288 specimens that were enrolled but were not tested on either Aptima SARS-CoV-2 or MAX SARS-CoV-2 since the positive target goal was reached.

**Supplemental Table 2.** Demographic information for compliant specimens with reportable Aptima SARS-CoV-2 and MAX SARS-CoV-2 results.

|  | **Characteristics** | **% (n/N)** |
| --- | --- | --- |
| **Gender** | Female | 59.6% (422/708) |
|  | Male | 40.4% (286/708) |
| **Age Group** | <18 Years | 4.2% (30/708) |
|  | 18 – 64 Years | 79.7% (564/708) |
|  | ≥65 Years | 15.8% (112/708) |
|  | Unknown | 0.3% (2/708) |

**Figure S1.**
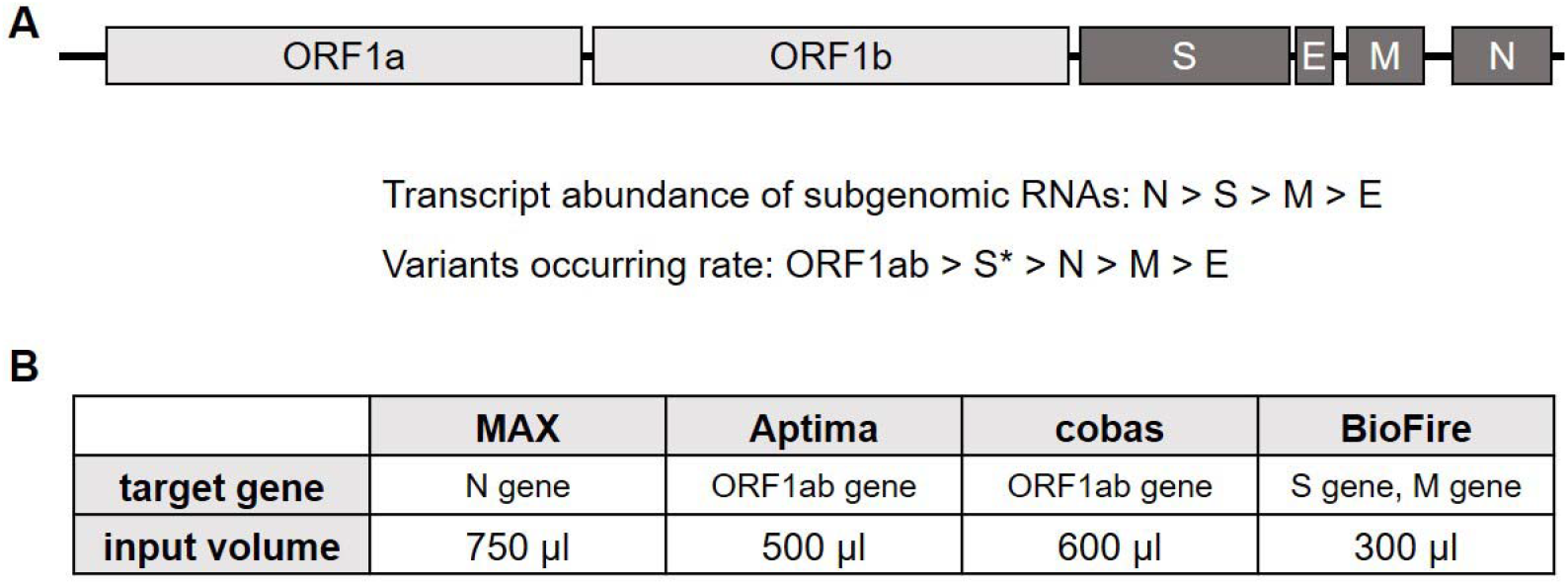
**(A)** Illustration of the genomic structure of SARS-CoV-2 gene based on the MN908497 Wuhan-Hu-1 sequence. Light gray domains are non-structural regions whereas dark gray domains code structural proteins. The order of the transcript abundance of subgenomic RNAs and variants occurring rate on each region are indicated.(14, 18) *Note that the mutations occur on S gene often impact the transmission of the virus. ORF: open reading frame; S: spike protein; E: envelope; M: membrane protein; N: nucleocapsid. **(B)** Summary of the targeted SARS-CoV-2 gene domain and minimum input volume for each RT-PCR system.(10, 19-21)

